# Evaluating Feature Selection Methods and Feature Contributions for Cardiovascular Disease Risk Prediction

**DOI:** 10.1101/2025.07.12.25331445

**Authors:** Suraiya Akhter, John H. Miller

**Affiliations:** School of Business and Technology, Emporia State University, Emporia, Kansas, USA; School of Engineering and Applied Sciences, Washington State University, Richland, Washington, USA

**Keywords:** Cardiovascular disease risk prediction, feature selection, machine learning, Shapley additive explanations, web application

## Abstract

**Background:** Cardiovascular disease (CVD) remains the foremost contributor to global illness and death, underscoring the critical need for effective tools that can predict risk at early stages to support preventive care and timely clinical decisions. With the growing complexity of healthcare data, machine learning has shown considerable promise in extracting insights that enhance medical decision-making. Nonetheless, the effectiveness and clarity of machine learning models largely rely on the relevance and quality of input features.

**Methods:** In this work, we explored and compared three distinct feature selection strategies—Alternating Decision Tree (ADT)-based analysis, Cross-Validated Feature Evaluation (CVFE), and Hypergraph-Based Feature Evaluation (HFE)—to isolate the most predictive clinical variables for assessing CVD risk. Our analysis utilized data from the National Health and Nutrition Examination Survey (NHANES), administered by the National Center for Health Statistics under the Centers for Disease Control and Prevention (CDC), encompassing demographic, clinical, laboratory, and survey data collected across the U.S. from August 2021 through August 2023. Distinct sets of features obtained through the selection techniques were used to develop eXtreme Gradient Boosting (XGBoost) models, which were then assessed for predictive effectiveness. To improve clarity and understand the model’s decision-making, SHapley Additive exPlanations (SHAP) was utilized to interpret the influence of each feature in the top-performing model.

**Results:** Among the approaches, the HFE method achieved the most accurate results, reaching 75% accuracy and an AUC of 0.7857, outperforming the alternatives. The most influential predictors identified by the best model included age, total cholesterol, glycohemoglobin levels, systolic blood pressure, smoking history, and a diagnosis of diabetes. The web application, accessible at https://shiny.tricities.wsu.edu/cvdr-prediction/, presents predictive results, probability scores, and a SHAP plot generated from the model trained using the feature set selected by the hypergraph-based approach.

**Conclusions:** This study highlights the importance of strategic feature selection in refining predictive accuracy and interpretability, offering a practical data-centric approach that could aid clinicians in evaluating cardiovascular risk and tailoring preventive care.

**Trial registration:** Not applicable as this research is not a clinical trial.

## Introduction

Globally, cardiovascular diseases (CVDs) remain the primary cause of death, responsible for around 17.9 million fatalities each year, which equates to nearly one-third of all deaths worldwide [1]. This broad category includes multiple heart and circulatory system disorders such as coronary artery disease, stroke, peripheral artery disease, rheumatic heart conditions, and congenital cardiovascular defects [2, 3]. Among these, coronary artery disease constitutes the majority, accounting for approximately 64% of CVD occurrences [2]. These statistics highlight the critical need for robust early detection methods and preventive healthcare measures to reduce the global impact of CVD. CVDs result from a multifaceted interaction between controllable and uncontrollable risk factors. Lifestyle-related elements such as elevated blood cholesterol, diabetes, excess body weight, tobacco use, and lack of physical activity are among the key modifiable influences [4, 5]. In contrast, unchangeable determinants include advancing age, biological sex, and racial or ethnic background [6–9]. The widespread adoption of unhealthy behaviors in today’s society has intensified these risks [10, 11]. Therefore, identifying high-risk individuals with precision and at an early stage is essential to enable preventive actions, slow disease progression, and decrease mortality rates.

Traditional clinical risk assessment tools—such as the Framingham Risk Score, SCORE risk charts, and the REGICOR risk score—have been widely used in practice [12–14]. However, these models often rely on a limited set of features and assume linear relationships between predictors and outcomes, potentially oversimplifying the true complexity of cardiovascular risk. Moreover, these models can be population-specific and may lack generalizability across diverse demographic and clinical settings. To address these challenges, machine learning has gained prominence as an advanced approach for improving the accuracy of cardiovascular risk assessment. Machine learning methods are capable of modeling complex, non-linear interactions among diverse features, offering greater flexibility and predictive accuracy than traditional statistical techniques [15–17]. Frequently applied machine learning algorithms in the prediction of cardiovascular diseases encompass decision tree models, support vector machines (SVM), logistic regression, *k*-nearest neighbors (KNN), random forest classifiers, gradient boosting methods, XGBoost, and advanced deep learning architectures like convolutional neural networks (CNNs) [2, 7, 18–22].

While personalized predictive modeling has advanced, significant difficulties remain in fully grasping the intricate relationships between various contributing factors, their differing effects across population subgroups, and how to tailor the most effective treatment plans for each individual. A broad range of socio-demographic, behavioral, and clinical factors contribute to variability in CVD outcomes, such as gender, age, race/Hispanic origin, education level, blood pressure readings, body mass index, waist circumference, lifestyle and physical activity indicators, sleep behavior, diabetes status, socioeconomic indicators, lipid profiles, glycemic control, inflammation markers, and smoking history. The complexity and interplay of these variables highlight the need for data-driven models capable of capturing these nuances. We hypothesize that a machine learning method can be harnessed to uncover and rank the most influential factors in predicting CVD risk, offering an objective and individualized framework for risk evaluation. Such a model has the potential to assist healthcare professionals in selecting the most suitable, patient-specific treatment plans to improve CVD outcomes.

For our study, we utilized data from the National Health and Nutrition Examination Survey (NHANES), a publicly accessible dataset curated by the Centers for Disease Control and Prevention (CDC). We developed a baseline predictive pipeline for CVD risk prediction using a machine learning algorithm and integrated robust feature selection techniques. By evaluating feature contributions, we aim to guide the collection and prioritization of essential clinical and lifestyle data for optimizing risk prediction and treatment decisions. This approach is intended to support evidence-based decision-making by health professionals and improve treatment outcomes for patients at risk of CVD. A web-based platform is available at https://shiny.tricities.wsu.edu/cvdr-prediction/, offering a comprehensive prediction framework that integrates the optimal feature evaluation method—hypergraph-based feature evaluation (HFE). The system supports SHAP-based interpretability and enables batch analysis of multiple samples. Users can also incorporate new data to further enhance the performance of the underlying predictive models.

## Materials and methods

Figure 1 outlines the full workflow of our proposed methodology. The process begins with collecting data from individuals classified as either at risk for CVD or not at risk. From this data, a pool of candidate features is generated. We then apply multiple feature evaluation techniques—including the Alternating Decision Tree (ADT) [23, 24], Cross-Validated Feature Evaluation (CVFE) [25], and Hypergraph-Based Feature Evaluation (HFE) [26]—to eliminate features with low relevance or minimal impact. The refined feature subsets are subsequently utilized to train an XGBoost model, which is evaluated for its predictive performance.

**Figure 1.**
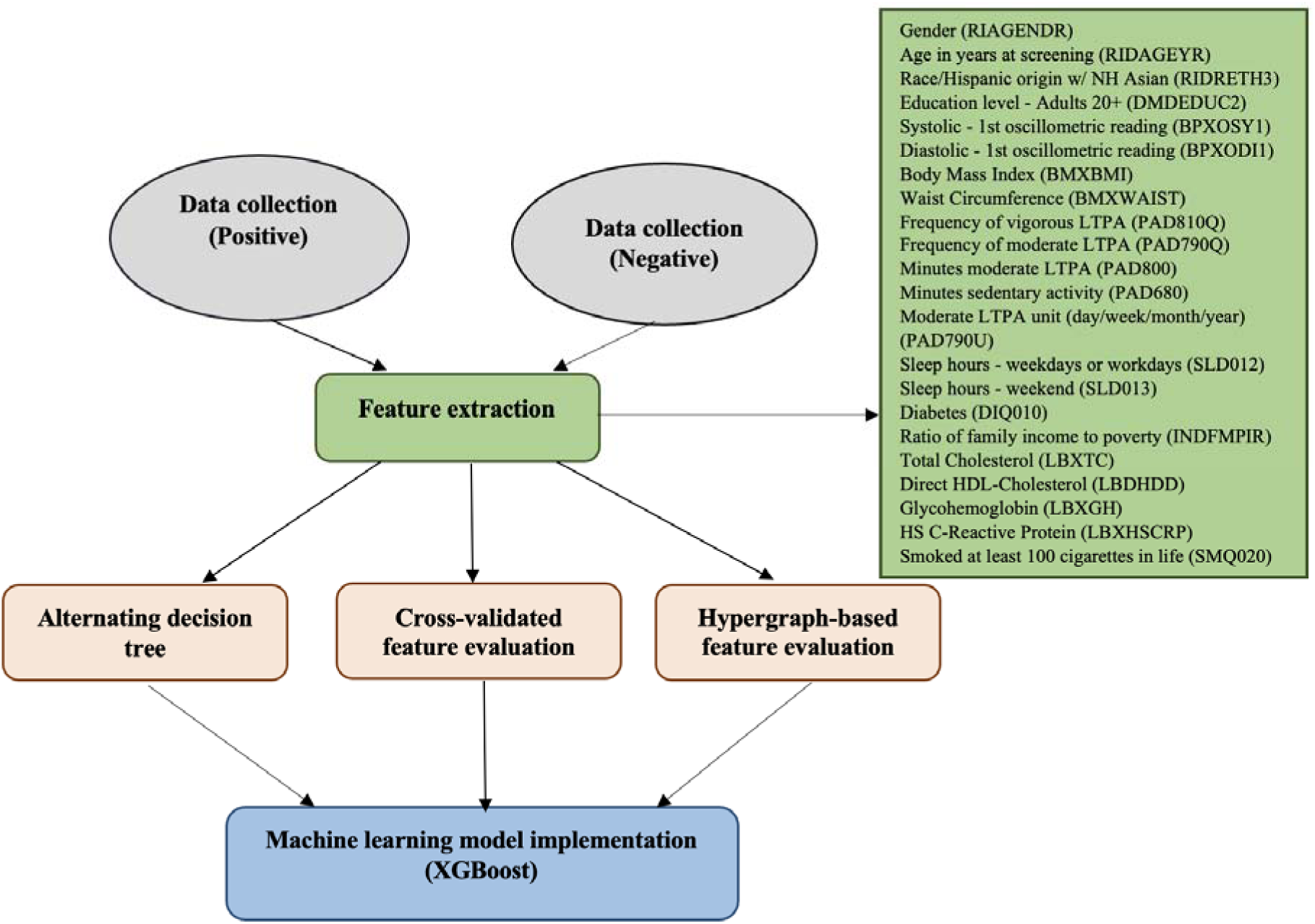
Overview of the procedure for detecting cardiovascular risk.

### Data and features

The data for this study was sourced from the NHANES, a publicly available program overseen by the National Center for Health Statistics and the Centers for Disease Control and Prevention (CDC) [27]. The final dataset consisted of 341 individuals identified as having CVD risk and 3,230 individuals without such risk. To mitigate class imbalance, a random sampling strategy was applied to downsample the non-CVD group to 341 cases, achieving a balanced dataset. To construct the model, the dataset was partitioned such that 80% was used for training purposes, while the remaining 20 was reserved for evaluation on a test set. Table 1 presents the selected variables, their categorical groupings, demographic distributions, and corresponding *p*-values, with a threshold of *p*□<□0.05 denoting statistical significance. The outcome variable, CVD risk, was defined such that individuals with a reported history of stroke, heart attack, coronary heart disease, or heart failure were classified as at risk for CVD. A total of 22 features were initially included as candidate predictors and are listed in Supplementary Table S1 (**Supplementary Material**).

**Table 1.**
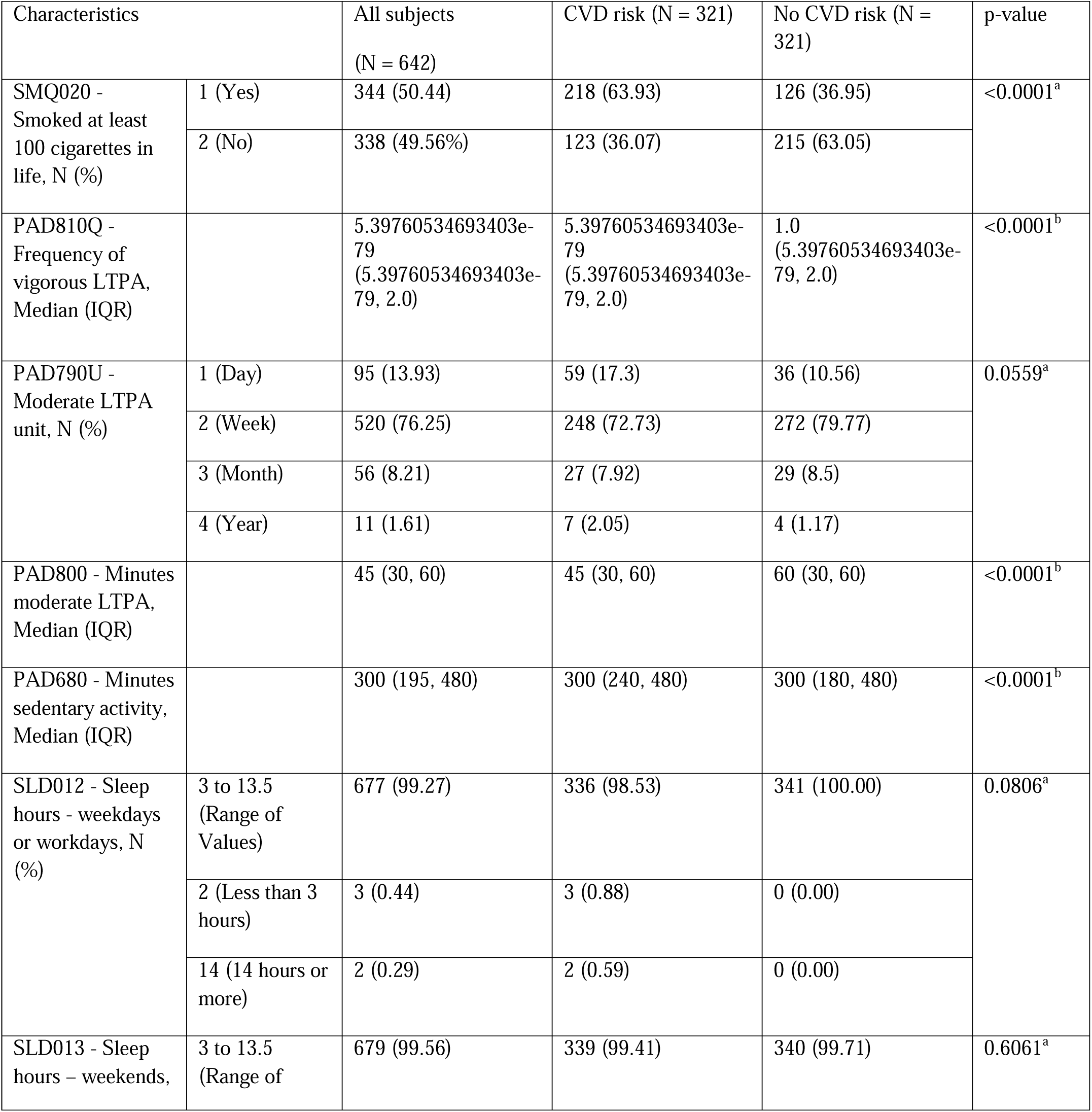
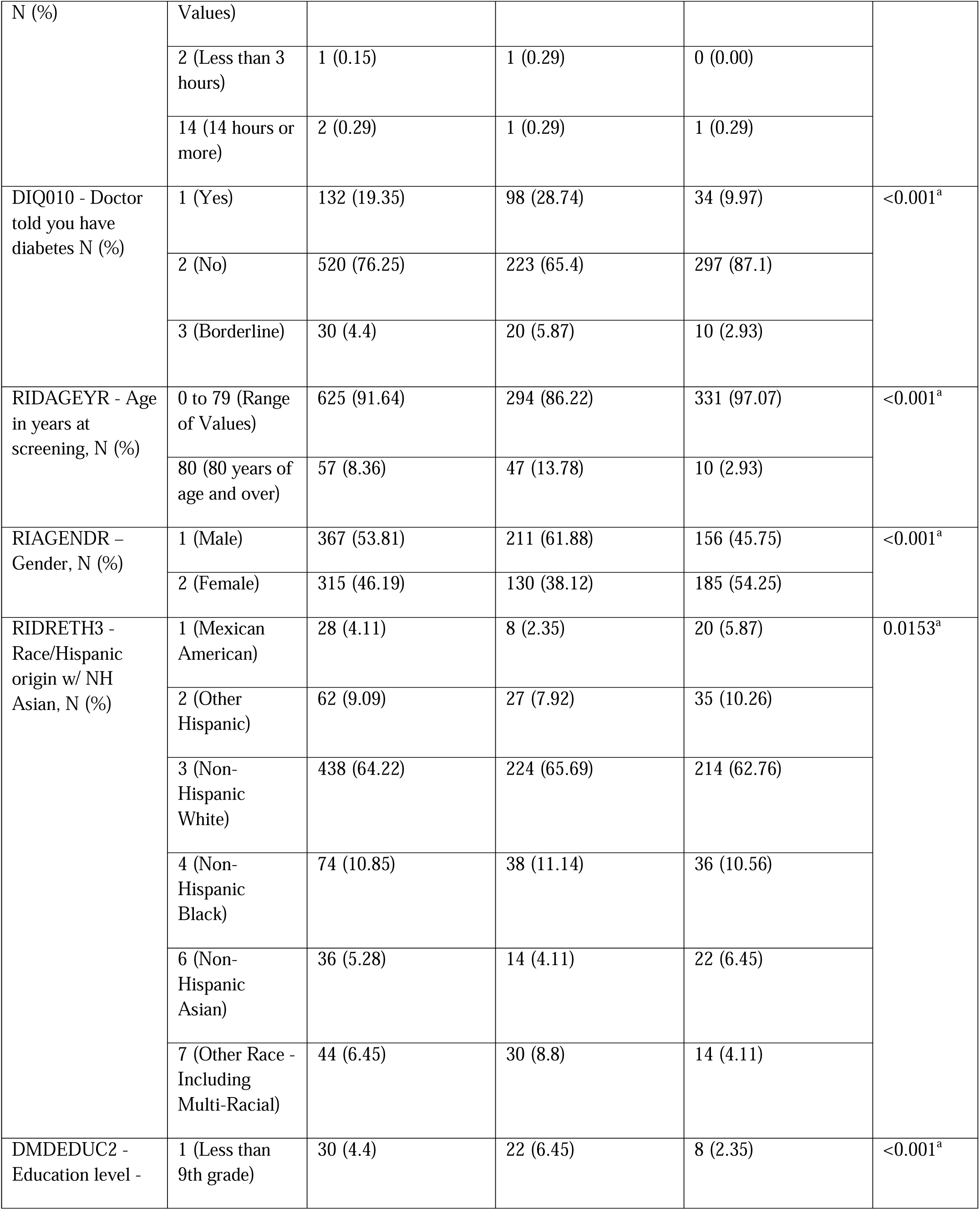
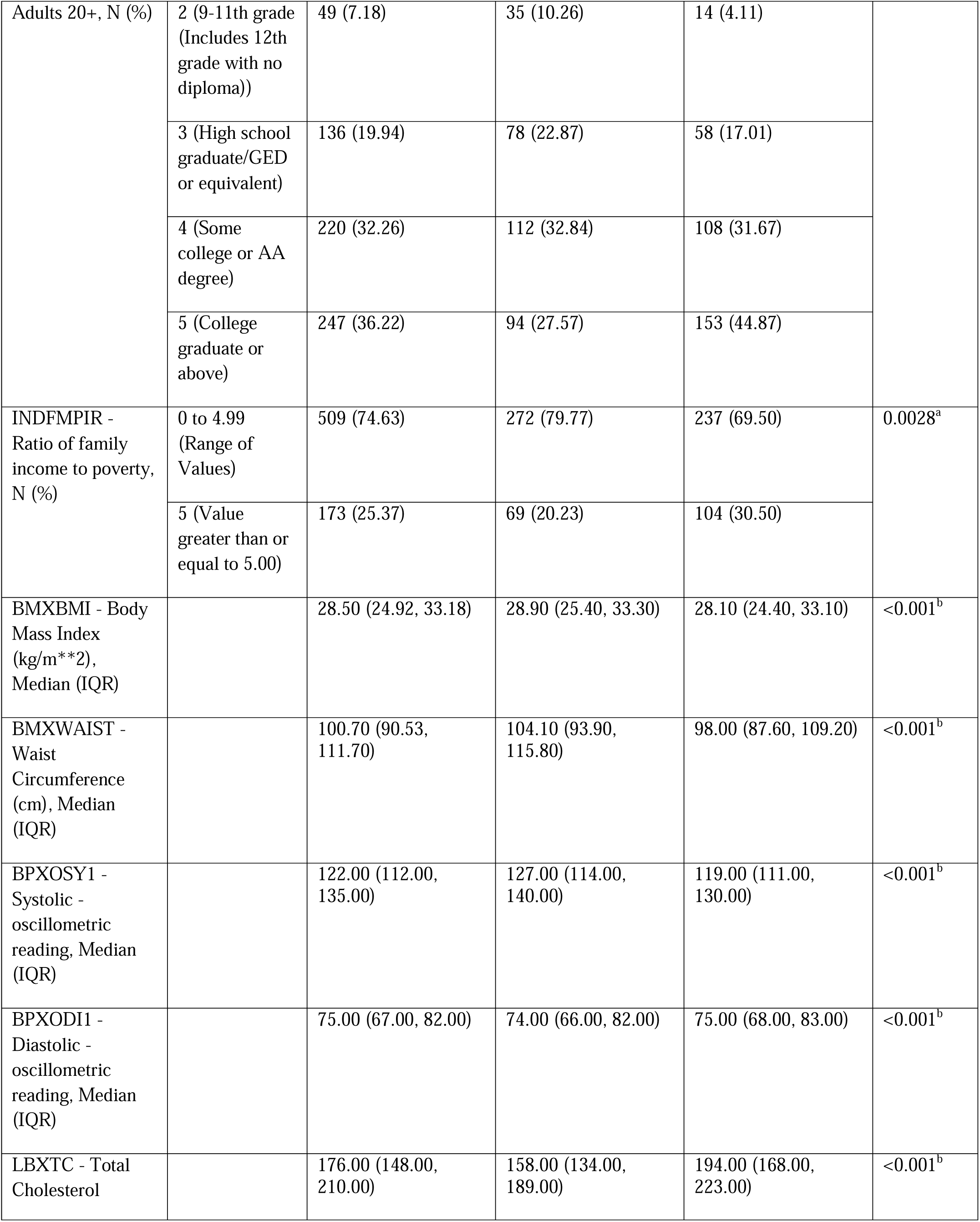
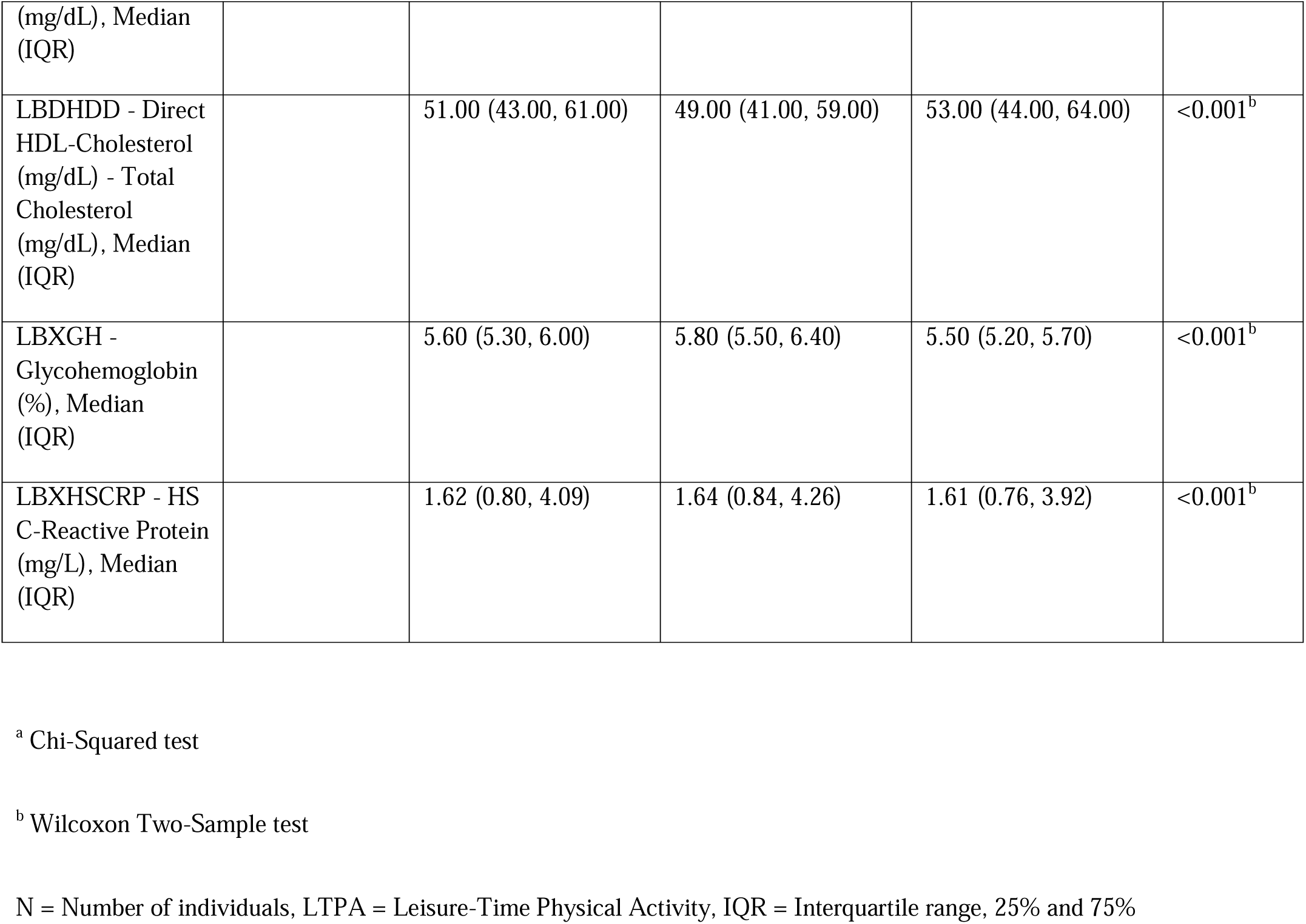
Summary of the demographic profile and clinical features of the study population.

### Assessment of predictive features

To build an effective predictive model, it is vital to discard non-contributory features during the preprocessing stage. In our study, we independently applied three feature reduction techniques—ADT [23, 24], CVFE [25], and HFE [26]—to refine the original feature set by eliminating attributes with limited predictive value. The ADT approach merges the straightforward, interpretable form of a conventional decision tree with the performance enhancements derived from boosting algorithms. This technique structures its model using decision tree stumps, which are foundational units commonly associated with boosting. One of the notable advantages of ADT is its flexible branching structure; unlike traditional trees with mutually exclusive paths, ADT allows overlapping routes, enabling multiple decision paths to contribute simultaneously to a prediction. The structure begins with a prediction node that assigns a numerical score, followed by layers of decision nodes that contain conditions used to evaluate input features. These layers alternate in a pattern—prediction nodes followed by decision nodes and vice versa. Decision nodes apply specific logical criteria, while prediction nodes assign fixed numeric contributions to the outcome. Importantly, prediction nodes appear at both the starting point (root) and terminal ends (leaves) of the tree, underscoring the distinctive, layered composition and operational logic that sets ADT apart from conventional decision tree models.

The ADT constructs a series of classification rules, each composed of three main elements: a prerequisite condition, a logical condition, and a pair of numerical scores. The logical condition takes the form of a predicate expressed as “feature <operator> threshold,” while the prerequisite is a compound logical statement formed by combining multiple such conditions using conjunctions. These rules are evaluated hierarchically using nested “if” statements, and their respective scores are used to compute the final prediction for a data sample. The procedure initiates with a root rule defined by unconditional logic— both the prerequisite and condition are set to “true”—and corresponding scores are computed using the weights assigned to training instances. Initially, each training sample is assigned an equal weight of 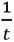, where *t* denotes the total number of training examples. During the training process, the ADT algorithm repeatedly generates new rules by identifying the most effective pair of prerequisite and condition that minimizes a specific objective function, denoted as *z*. This function evaluates the discriminatory power of a rule based on its ability to separate positive and negative classes effectively. For every new rule created, updated scores are computed through a boosting-based mechanism. Training sample weights are also revised in accordance with the rule’s classification accuracy on each example, emphasizing incorrectly predicted instances to refine future splits. This iterative process continues until a predetermined stopping criterion is satisfied—such as reaching the maximum number of iterations or when further performance gains become negligible. The resulting rule set defines an alternating decision tree, where each prediction node holds a scalar value, and the tree’s topology is dictated by the prerequisite logic embedded within the constructed rules. Only a portion of the total feature set is used in the final ADT, reflecting the most relevant variables identified through this process. For implementation, we evaluated 50 randomly selected values for *B*, representing the number of boosting cycles. The final ADT model—shown in Supplementary Figure S1 (**Supplementary Material**)—demonstrates the decision structure derived from 22 candidate features. Ultimately, 14 features were retained through the ADT-based selection process, as presented in Supplementary Table S2 (**Supplementary Material**).

In addition, we incorporated both the CVFE approach and a hypergraph-based technique to refine the initial collection of features. The CVFE process is illustrated in Figure 2. Initially, the dataset was randomly divided into *c* distinct subsets. For each subset, we applied the XGBoost algorithm to determine the most influential features, optimizing model hyperparameters through a grid search procedure. Feature selection was conducted independently for every subset, followed by the construction of an intersected feature set comprising features common to all subsets. This entire process was repeated *e* times, resulting in *e* intersected feature sets. Subsequently, any feature appearing in at least (*p* × 100)% of these intersected sets was incorporated into the final selected feature list. Table 2 details the quantity of features obtained from CVFE under various parameter settings of *c*, *e*, and *p*. A comprehensive list of the selected key features can be found in Supplementary Tables S3-S6 included in the **Supplementary Material**.

**Figure 2.**
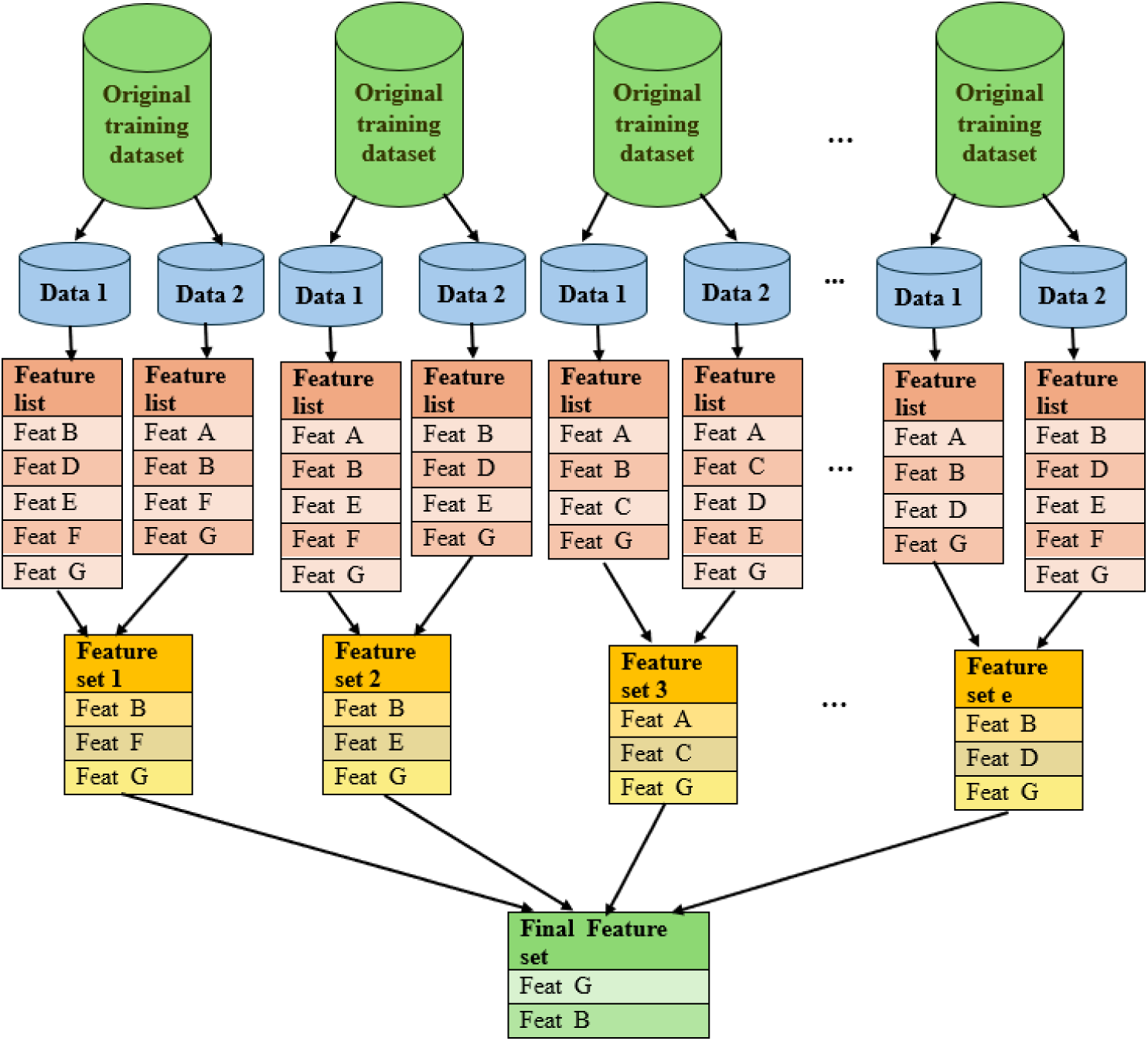
Visual representation of the feature selection process using the Cross-Validated Feature Evaluation (CVFE) method.

**Table 2.**
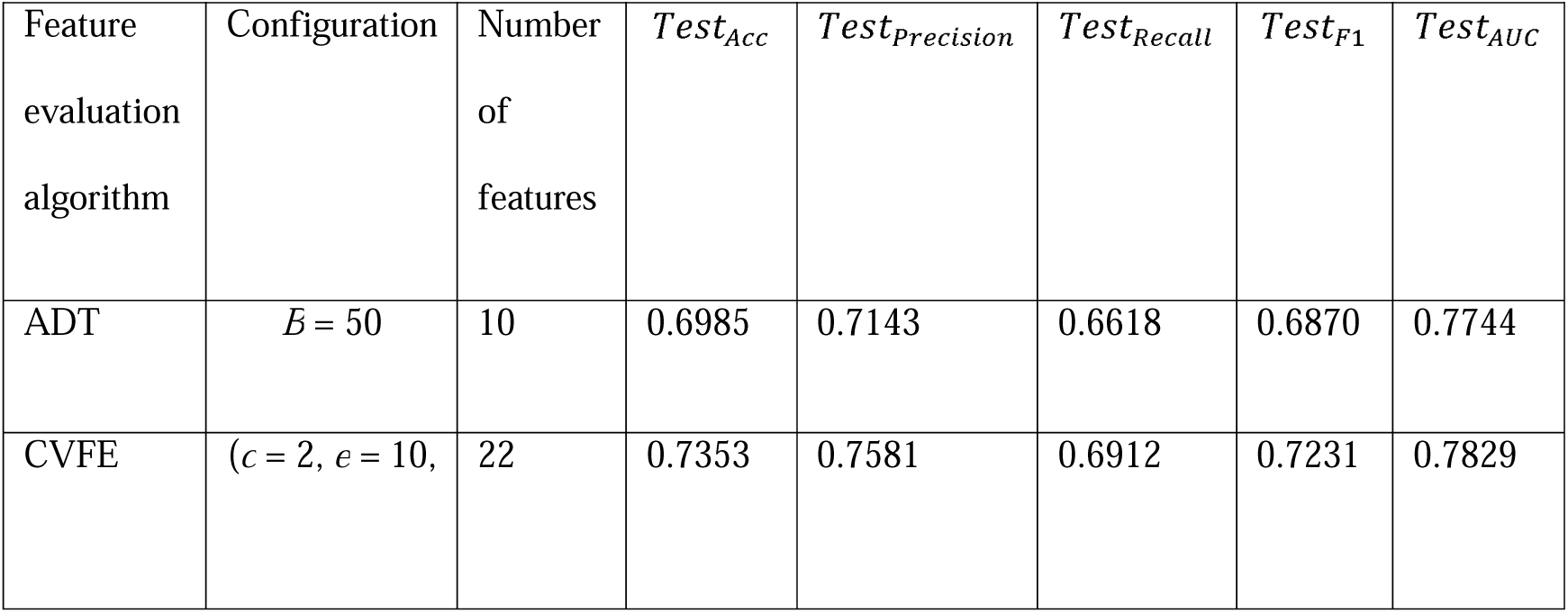
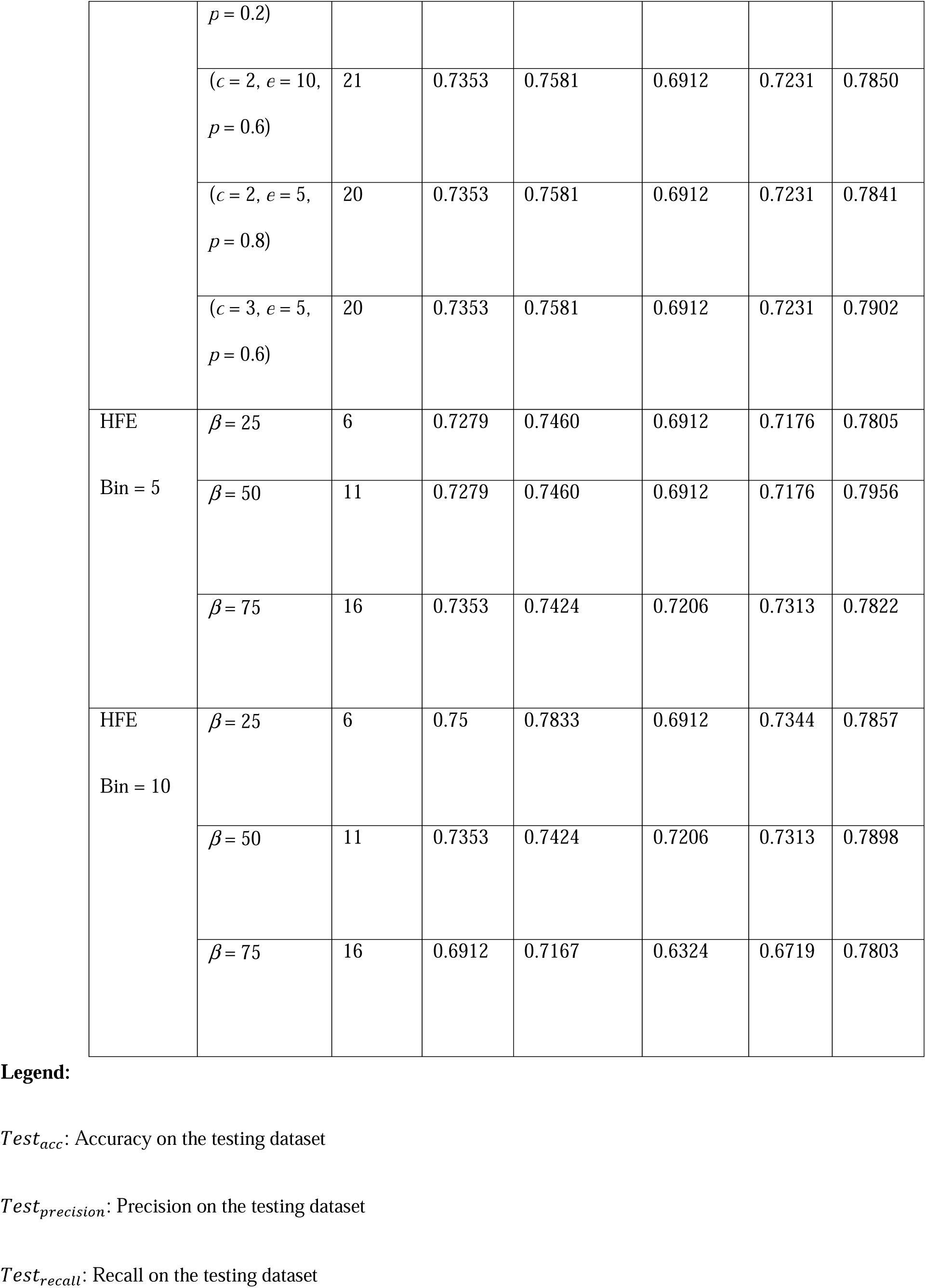
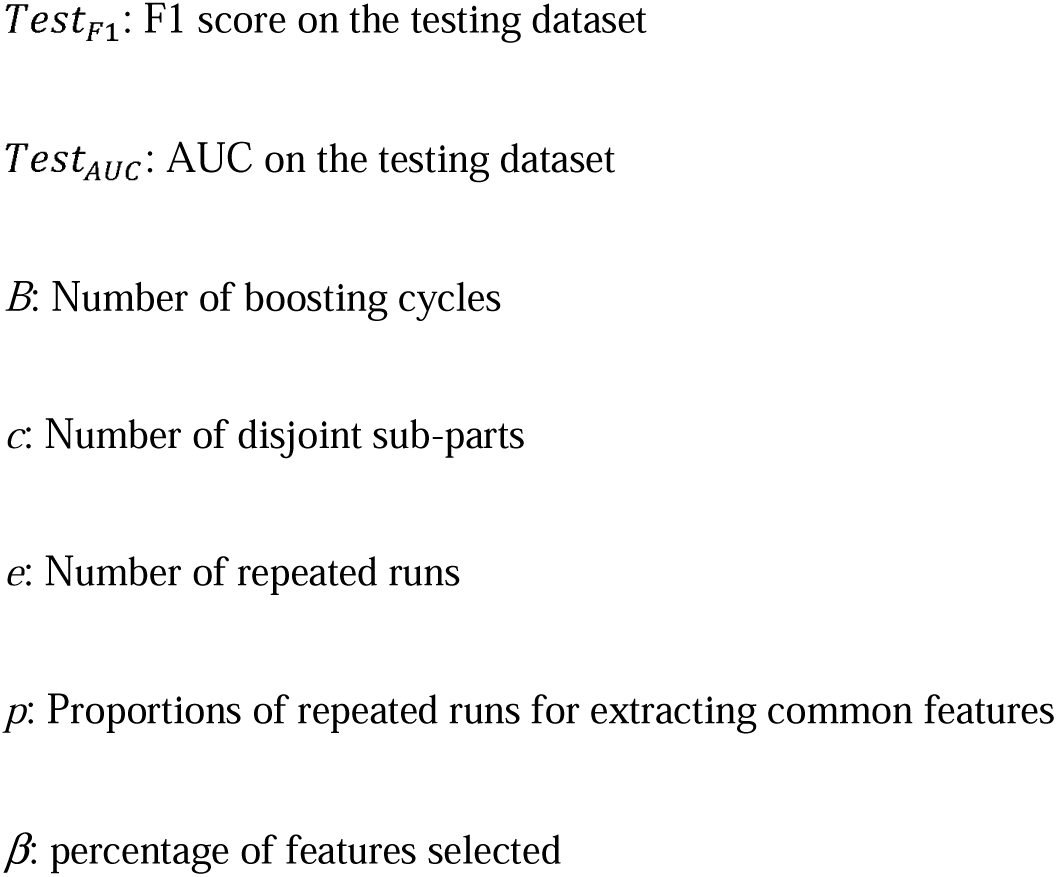
Summary of the number of selected features and corresponding evaluation metrics—accuracy, precision, recall, F1 score, and AUC—on the testing dataset for different feature subsets.

The HFE technique involves modeling feature interactions through a hypergraph structure. Unlike traditional graphs, represented as G(*V*, *E*), where vertices (*V*) are connected by pairwise edges (*E*), a hypergraph generalizes this concept by permitting each edge—termed a hyperedge—to link multiple vertices simultaneously. Formally, a hypergraph is defined as G(*V*, *E*), with *V* as the set of nodes and *E* comprising hyperedges, each of which is a subset of V. Figure 3 contrasts a standard graph with a hypergraph, while a hypergraph visualization of the complete training dataset is provid d in Supplementary Figure S2 (**Supplementary Material**). In applying HFE, each continuous feature was discretized into a fixed number of intervals (bins), and the feature relevance scores were computed based on the complex, multi-dimensional relationships captured within the hypergraph structure. These scores were derived by assigning weights to the hyperedges that represent the corresponding feature bins. The ranking of features was guided by the hypergraph cut conductance minimization framework [26], ultimately producing a rating vector that reflects the relative importance of each feature. To determine the final feature subset, the top *z* features were selected based on their sorted importance values, where z is computed as *β* × *m* (*m* being the total number of features and *β* representing the selection ratio). In our experiments, we used discretization settings of bin = 5 and bin = 10. The resulting feature counts for varying *β* values and bin sizes are reported in Table 2. The specific features retained through HFE under each bin configuration are detailed in Supplementary Tables S6 and S7 (**Supplementary Material**).

**Figure 3.**
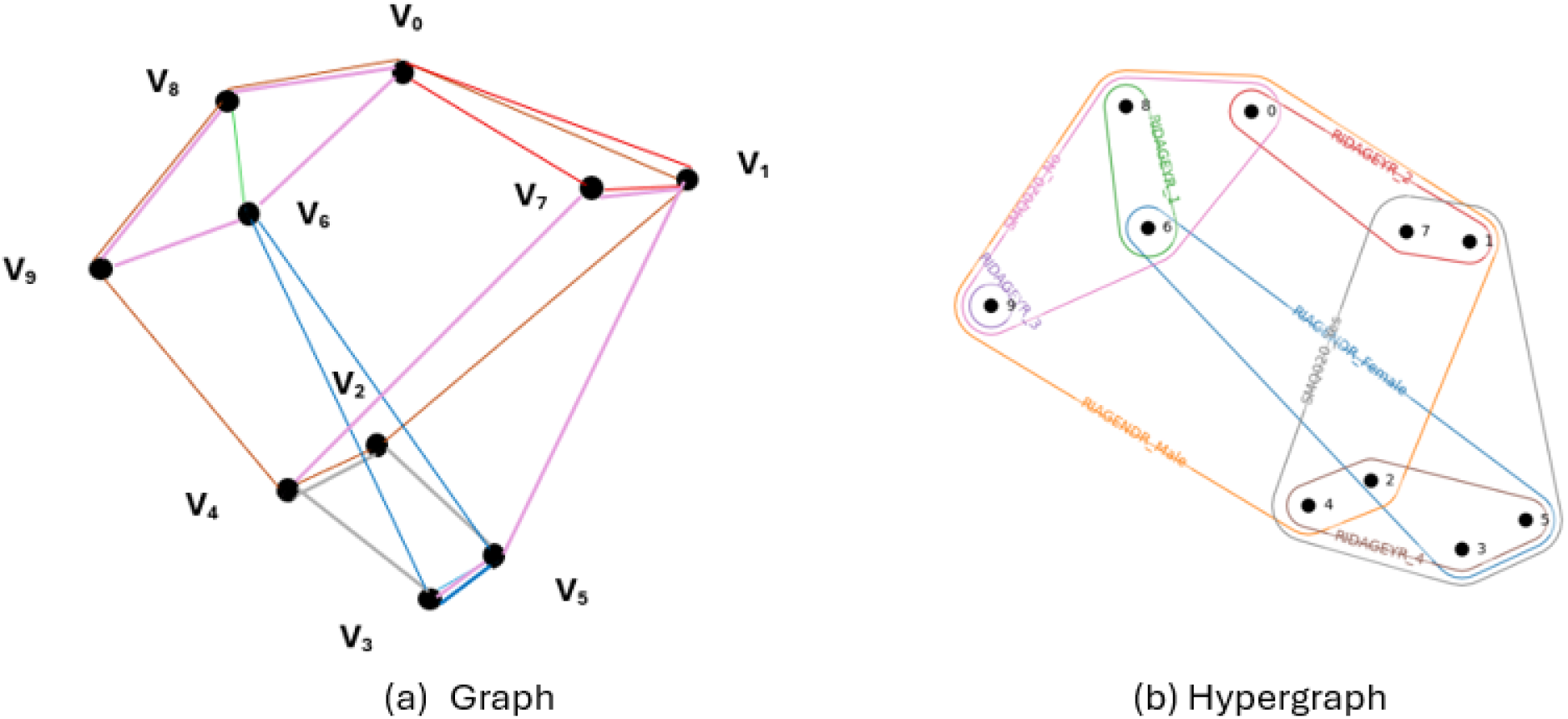
Comparison between a standard graph and a hypergraph.

### Web application

As shown in Figure 4, the HFE approach was incorporated into a machine learning-powered web application. This tool delivers both classification outcomes and associated probability scores. Instructions for uploading datasets, performing binary classification, and estimating probabilities are provided within the application interface. Users have the option to download relevant output files and contribute additional training data, thereby improving model performance. The tool also supports the export of a SHAP visualization, allowing users to examine how top-ranked features influence predictions. The web application is publicly available at https://shiny.tricities.wsu.edu/cvdr-prediction/.

**Figure 4.**
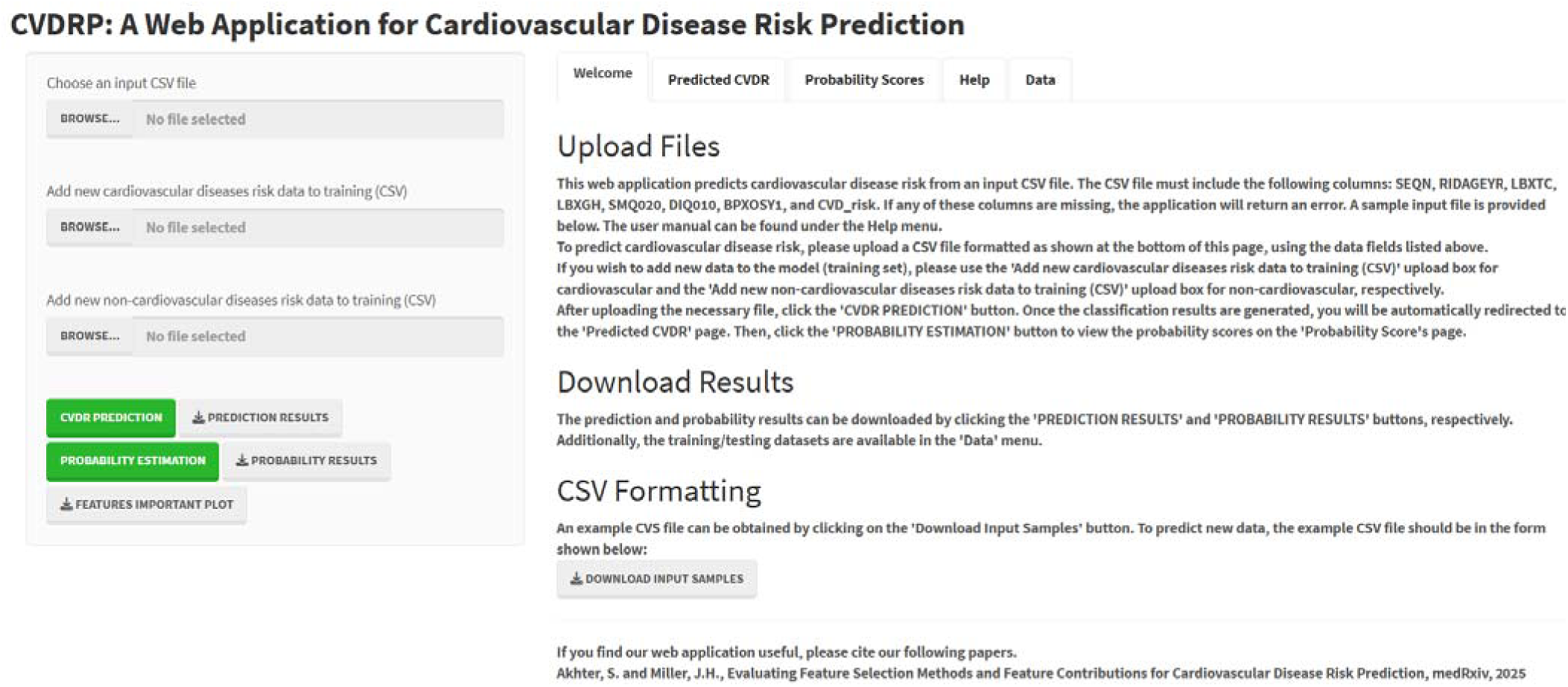
The web application for cardiovascular disease risk prediction.

### Code and data availability

The full set of experimental data along with all relevant scripts can be accessed at https://github.com/suraiya14/CVDRP, as referenced in the **Supplementary Material**.

## Results

Following the reduction of the original feature set using three independent feature selection strategies, we developed individual predictive models based on the selected features using the XGBoost algorithm [28]. To interpret and analyze the contribution of each feature within these models, we applied SHapley Additive exPlanations (SHAP) [29]. SHAP provides a unified framework for attributing the impact of each input variable by calculating how much each feature contributes to the model’s output, effectively measuring its marginal effect on prediction outcomes.

### Performance assessment

An XGBoost-based predictive model was constructed by utilizing different sets of features selected through ADT, CVFE, and HFE methodologies in conjunction with the training data. The model’s performance was assessed on the test data using the metrics defined in Equations 1 - 4, where TP, TN, FP, and FN represent true positive, true negative, false positive, and false negative counts, respectively. Among these evaluation metrics, accuracy was employed to determine the ratio of correctly predicted outcomes relative to the overall number of samples in the dataset.

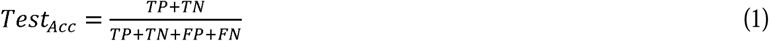

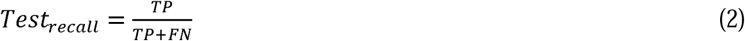

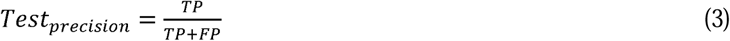

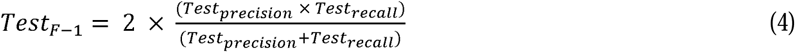

In addition, we computed recall and precision to further evaluate model performance. Recall quantifies the ability of the model to correctly detect actual positive cases, whereas precision reflects the proportion of positive predictions that are genuinely correct. To provide a balanced evaluation that incorporates both precision and recall, we utilized the F1 score, which is defined as their harmonic mean. Moreover, the Area Under the Curve (AUC) metric was used to assess the effectiveness of the binary classifier. Higher AUC values correspond to stronger model performance, with a score of 1 indicating flawless classification and 0.5 representing performance equivalent to random guessing.

Table 2 presents a detailed comparison of XGBoost model performance using feature subsets selected through ADT, CVFE, and HFE techniques. Corresponding confusion matrices for each reduced feature set are visualized in Supplementary Figure S3 (**Supplementary Material**). Overall, the HFE feature set (bin=10, *β*=25) yielded superior predictive performance compared to the ADT and CVFE sets, with the best model successfully identifying 47 out of 60 patients at risk for cardiovascular disease.

### Features identified using the HFE Approach

The highest predictive performance was attained using the XGBoost algorithm paired with the HFE feature selection method configured with bin=10 and *β*=25. Figure 5 presents a SHAP summary bar plot illustrating the importance rankings of the features identified through SHAP value analysis applied to that model. From an initial set of 22 features, HFE selected 6 key predictors: RIDAGEYR (age at screening), LBXTC (total cholesterol in mg/dL), LBXGH (glycohemoglobin), BPXOSY1 (systolic blood pressure via oscillometric measurement), SMQ020 (history of smoking at least 100 cigarettes), and DIQ010 (physician-diagnosed diabetes), ordered according to their relative influence on the model’s predictions.

**Figure 5.**
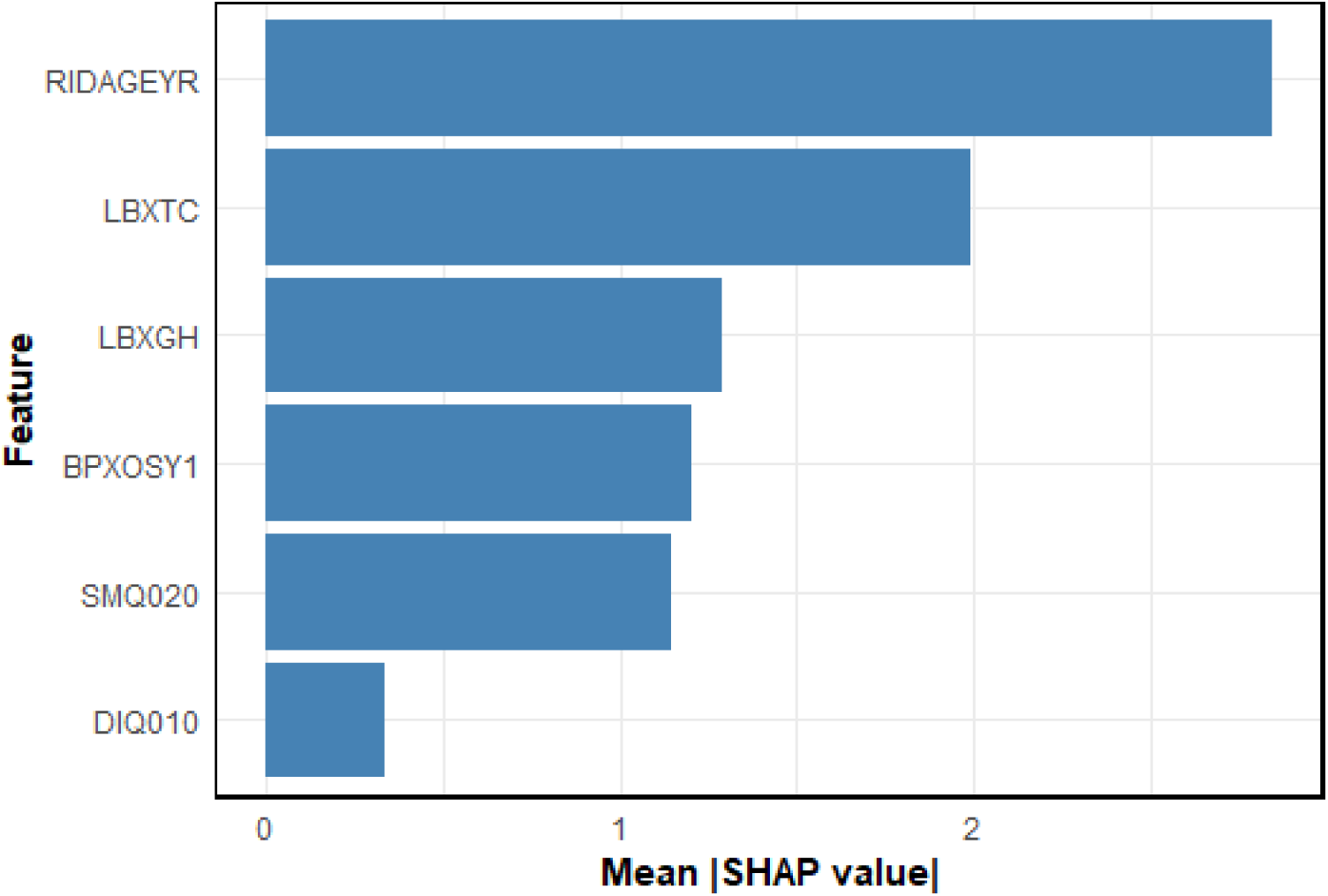
Importance of features selected by the XGBoost model shown as a bar chart. The *x*-axis represents the mean absolute SHAP value for each feature, reflecting its average contribution to the model’s predictions. Features are arranged from most to least influential.

### Feature impact assessment

Figure 6 presents the importance of the key continuous features identified in Figure 5 through SHAP value analysis. Each point corresponds to an individual patient, with overlapping points slightly offset (jittered) to reflect the density of data. The *x*-axis displays how much each feature influences the model’s prediction, where higher SHAP values correspond to an elevated risk of CVD. The color gradient—from yellow to purple—represents low to high feature values, respectively. Notably, increased SHAP values for age, glycohemoglobin levels, and systolic blood pressure (oscillometric measurement) are linked to a greater likelihood of CVD. While higher total cholesterol often aligns with increased risk, this pattern does not hold consistently across all patients.

**Figure 6.**
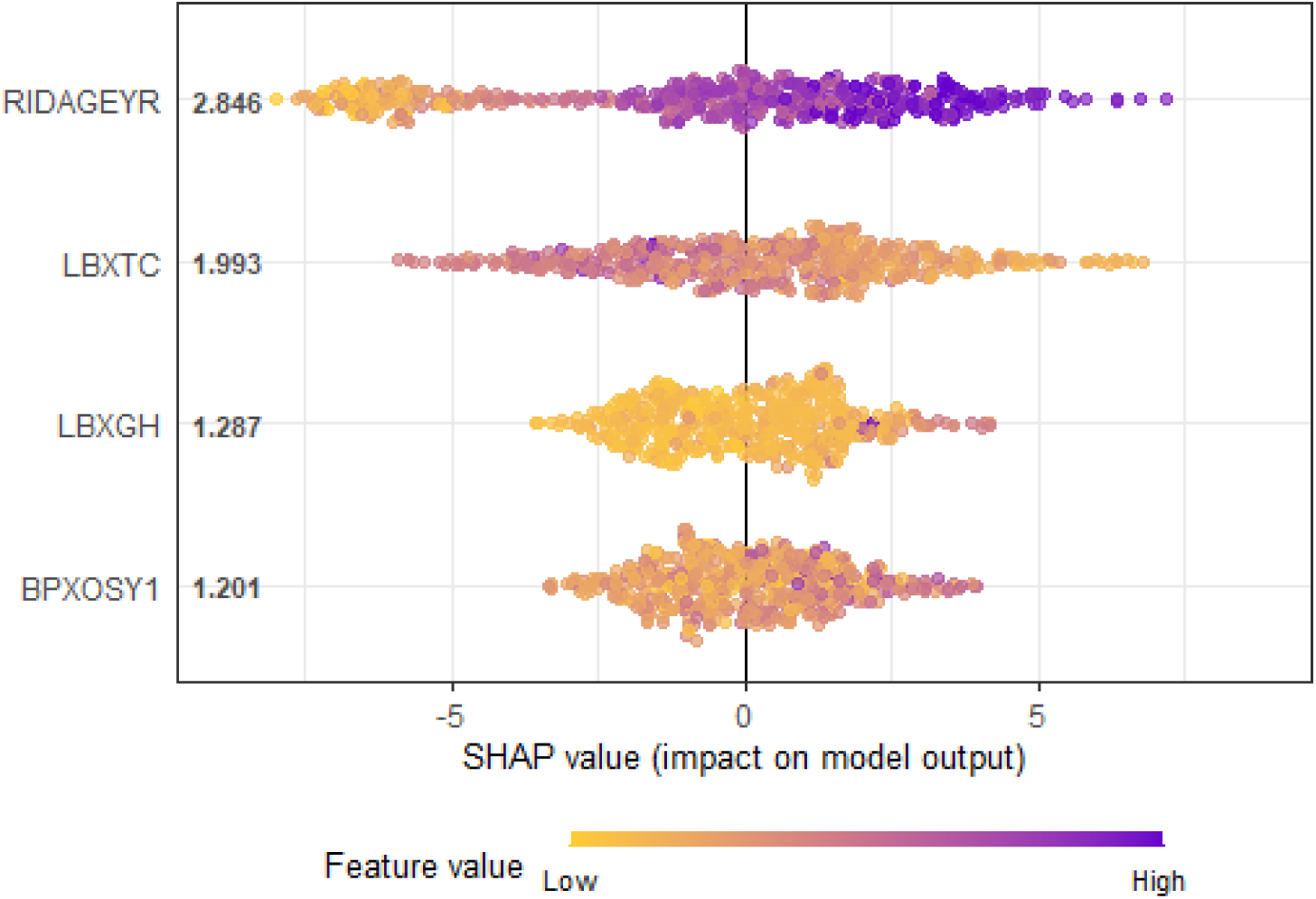
SHAP analysis illustrating the impact of the top continuous features on model predictions.

Figure 7 displays boxplots representing the SHAP values for the different categories of the categorical features identified in Figure 5. Since elevated SHAP values correspond to increased CVD risk, the boxplots allow for straightforward interpretation of risk levels across categories. Specifically, Figure 7(a) shows that individuals who have smoked at least 100 cigarettes in their lifetime exhibit a higher CVD risk. Likewise, Figure 7(b) indicates that patients diagnosed with diabetes tend to have an elevated risk of developing CVD.

**Figure 7.**
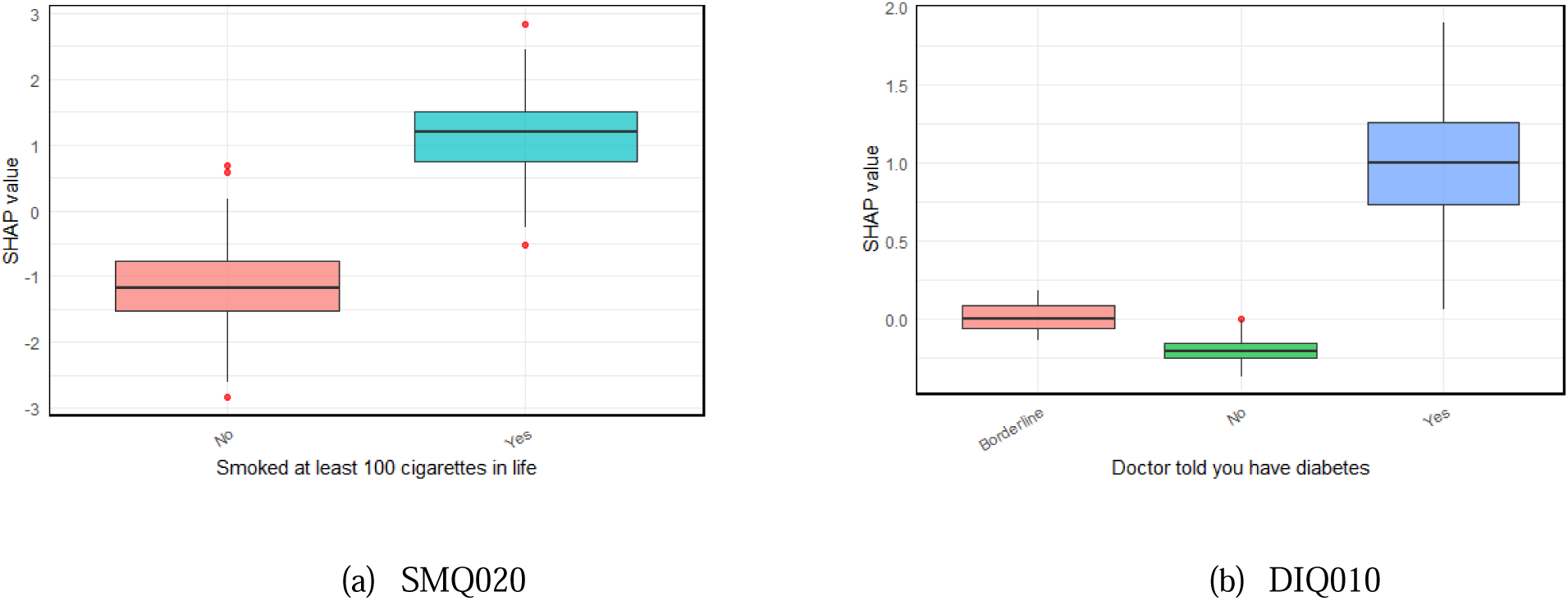
SHAP analysis illustrating the impact of the top categorical features on model predictions.

## Discussion and conclusions

Accurate individualized prediction plays a vital role in enhancing patient outcomes in CVD risk evaluation. This work introduced a CVD risk prediction model built on the XGBoost machine learning framework. By systematically removing unnecessary and non-contributory features, we assessed the model’s effectiveness using a streamlined feature subset. The resulting web application incorporates the hypergraph-based strategy and enables users to apply this method to predict cardiovascular risk in external test datasets. Furthermore, users can expand the training dataset by incorporating additional samples and conduct SHAP-based interpretation to explore the influence of key features. Therefore, the prediction pipeline successfully pinpointed critical variables affecting CVD risk, offering valuable insights for tailored risk assessment, guiding therapeutic approaches, and aiding clinical decision processes.

Our results are consistent with existing literature. For example, Peng et al. [30] demonstrated that the XGBoost and XGBH algorithms achieved superior predictive accuracy, with systolic blood pressure identified as the most influential variable. Likewise, Khaw and Wareham [31] emphasized the significant link between glycated hemoglobin levels and cardiovascular risk, noting that even slight increases below the diabetic threshold correspond to a 10–20% elevation in CVD risk. The widely implemented QRISK3 model in the UK incorporates factors such as age, systolic blood pressure, cholesterol ratios, smoking habits, and diabetes status as primary indicators of cardiovascular risk [32]. Additionally, Shishehbori and Awan [33] surveyed recent machine learning methods in this field, underscoring that conventional clinical markers— including age, blood pressure, cholesterol, smoking history, and diabetes—continue to be the most influential predictors across diverse models.

Our model highlighted age as the most impactful predictor, a conclusion corroborated by multiple prior investigations [34–37] that emphasize age as a key determinant of cardiovascular disease risk. Furthermore, the identification of other important variables—including total cholesterol, glycated hemoglobin, systolic blood pressure, smoking history, and diabetes status—carries substantial clinical relevance. Prioritizing these features allows healthcare providers to enhance the accuracy of risk stratification, tailor prevention efforts, and effectively allocate resources for managing cardiovascular conditions. While our methodology offers notable advantages, several limitations must be acknowledged. The accuracy of feature contribution assessments may be compromised in subpopulations where data are limited. Moreover, the present model is not designed to handle time-to-event analyses or longitudinal forecasting, restricting its usefulness for tracking disease progression over time. Future research should aim to expand the model’s capabilities to address these outcomes.

In conclusion, we constructed and validated a machine learning framework to estimate cardiovascular disease risk by leveraging a carefully selected set of features from NHANES datasets. Although the model achieved satisfactory predictive performance, there remains room for improvement in both accuracy and AUC metrics. The inclusion of more detailed, high dimensional data sources, such as multi-omics profiles or longitudinal clinical measurements, has the potential to boost predictive precision and clinical relevance.

## Data Availability

All data produced are available online at https://github.com/suraiya14/CVDRP.

## Supporting information

The supplementary material for this article is provided herewith the manuscript. (DOCX)

## Declarations

### Ethics approval and consent to participate

The study was conducted in accordance with the Declaration of Helsinki. The work is based on publicly available data; therefore, consent to participate was not required.

### Consent for publication

Consent for publication was obtained.

### Authors’ contributions

SA: Conceptualization, data collection, formal analysis, software implementation, validation, visualization, and writing manuscript. JHM: Conceptualization, supervision, reviewing analyses, and editing the manuscript. All authors read and approved the final manuscript.

## Acknowledgements

Not applicable.

