## Supplementary Materials for "Evaluating Feature Selection Methods and Feature Contributions for Cardiovascular Disease Risk Prediction"

### 1 Supplementary Data

#### 1.1 Training dataset

The training dataset can be accessible at https://github.com/suraiya14/CVDRP.

#### 1.2 Testing dataset

The testing dataset can be accessible at https://github.com/suraiya14/CVDRP.

### 2 Supplementary Tables

#### 2.1 Feature Set before applying feature reduction algorithm

**Table S1.** List of features used for feature reduction analysis

| SMQ020 | PAD790U | SLD012 | RIDAGEYR | DMDEDUC2 | BMXWAIST | LBXTC | LBXHSCRP |
| --- | --- | --- | --- | --- | --- | --- | --- |
| PAD810Q | PAD800 | SLD013 | RIAGENDR | INDFMPIR | BPXOSY1 | LBDHDD |  |
| PAD790Q | PAD680 | DIQ010 | RIDRETH3 | BMXBMI | BPXODI1 | LBXGH |  |

#### 2.2 ADT reduced features

**Table S2.** List of features obtained from ADT

| RIDAGEYR | PAD810Q | LBXTC | LBXHSCRP | BPXOSY1 |
| --- | --- | --- | --- | --- |
| SMQ020 | BMXWAIST | SLD012 | SLD013 | BPXODI1 |
| INDFMPIR | DMDEDUC2 | LBXGH | PAD680 |  |

#### 2.3 CVFE reduced features

In the following tables S3-S6, c, *e* and *p* indicate count of disjoint sub-parts, count of iterations and ratios of recurring iterations for the extraction of common features, respectively.

**2.3.1 Table S3.** List of features obtained from CVFE (*c* = 2, *e* = 10, *p* = 0.2)

| RIDAGEYR | SMQ020 | LBXTC | INDFMPIR | DMDEDUC2 | LBDHDD | SLD013 |
| --- | --- | --- | --- | --- | --- | --- |
| PAD810Q | PAD680 | SLD012 | BPXOSY1 | RIDRETH3 | PAD800 | PAD790Q |
| BMXBMI | DIQ010 | LBXGH | BMXWAIST | LBXHSCRP | BPXODI1 | PAD790U |
| RIAGENDR |  | | | | | |

**2.3.2 Table S4.** List of features obtained from CVFE (*c* = 2, *e* = 10, *p* = 0.6)

| RIDAGEYR | SMQ020 | LBXTC | INDFMPIR | DMDEDUC2 | LBDHDD | SLD013 |
| --- | --- | --- | --- | --- | --- | --- |
| PAD810Q | PAD680 | SLD012 | BPXOSY1 | RIDRETH3 | PAD800 | PAD790Q |
| BMXBMI | LBXGH | BMXWAIST | LBXHSCRP | BPXODI1 | PAD790U | RIAGENDR |

**2.3.3 Table S5.** List of features obtained from CVFE (*c* = 2, *e* = 5, *p* = 0.8)

| RIDAGEYR | SMQ020 | LBXTC | INDFMPIR | DMDEDUC2 | LBDHDD | SLD013 |
| --- | --- | --- | --- | --- | --- | --- |
| PAD810Q | PAD680 | SLD012 | BPXOSY1 | PAD800 | RIDRETH3 | PAD790Q |
| BMXBMI | LBXGH | BMXWAIST | LBXHSCRP | BPXODI1 | RIAGENDR |  |

**2.3.4 Table S6.** List of features obtained from CVFE (*c* = 3, *e* = 5, *p* = 0.6)

| RIDAGEYR | SMQ020 | LBXTC | INDFMPIR | DMDEDUC2 | LBDHDD | SLD013 |
| --- | --- | --- | --- | --- | --- | --- |
| PAD810Q | PAD680 | SLD012 | BPXOSY1 | PAD800 | PAD790Q | BMXBMI |
| RIDRETH3 | LBXGH | BMXWAIST | LBXHSCRP | BPXODI1 | RIAGENDR |  |

#### 2.4 HFE reduced features

The lists of features obtained from the HFE method—using bin values of 5 and 10 to discretize each feature—are presented in Tables S7 and S8. In each table, the first 6, 11, and 17 features correspond to β values of 25, 50, and 75, respectively.

**2.4.1 Table S6.** List of features obtained from HFE with bin = 5

| RIDAGEYR | SMQ020 | DIQ010 | LBXTC | DMDEDUC2 | SLD012 | RIAGENDR | SLD013 |
| --- | --- | --- | --- | --- | --- | --- | --- |
| LBXGH | INDFMPIR | BPXOSY1 | BMXWAIST | LBDHDD | RIDRETH3 | PAD790U | PAD680 |

**2.4.2 Table S7.** List of features obtained from HFE with bin = 10.

| RIDAGEYR | LBXTC | LBXGH | SMQ020 | DIQ010 | BPXOSY1 | DMDEDUC2 | SLD012 |
| --- | --- | --- | --- | --- | --- | --- | --- |
| RIAGENDR | SLD013 | LBDHDD | INDFMPIR | BMXWAIST | PAD680 | BPXODI1 | RIDRETH3 |

**3. Supplementary Figures**

**3.1 ADT**

**Figure S1 –** An alternative decision tree representation of the reduced feature set.

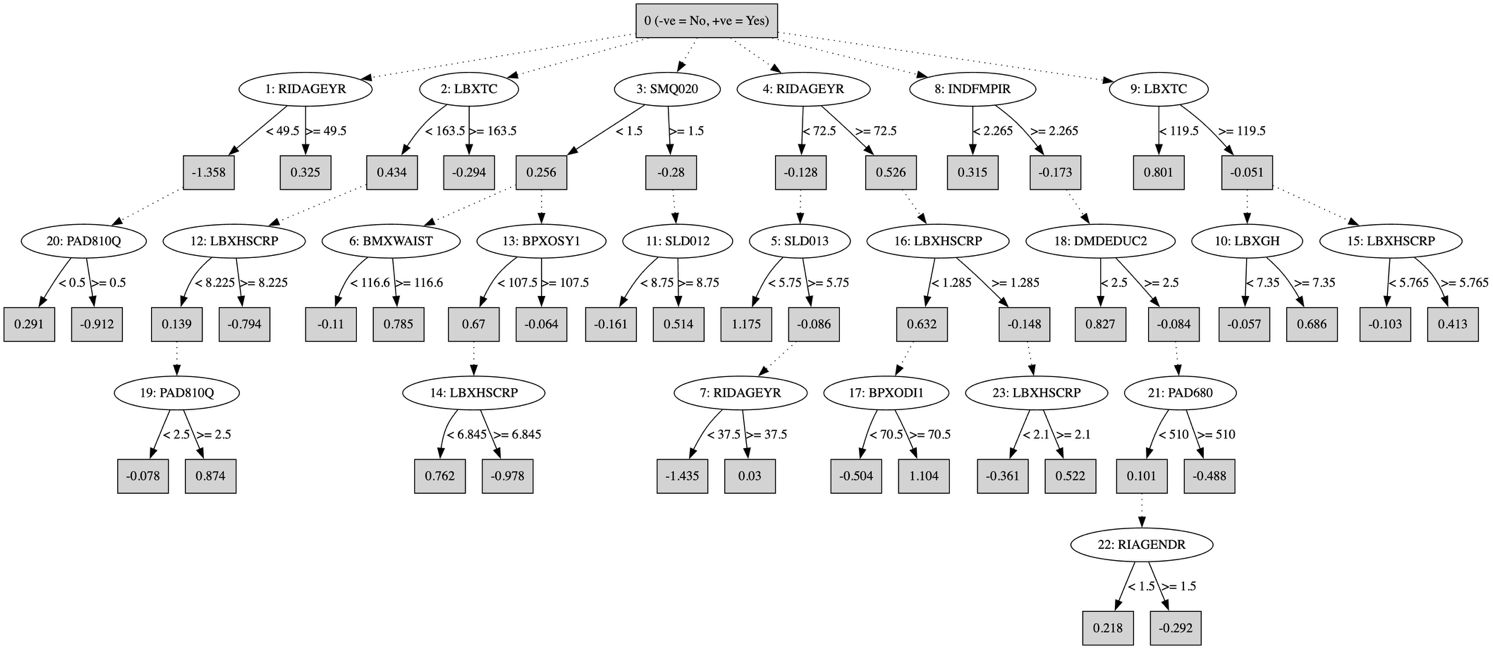

**3.2 Hypergraph**

**Figure S2 –** Hypergraph representation of the training dataset

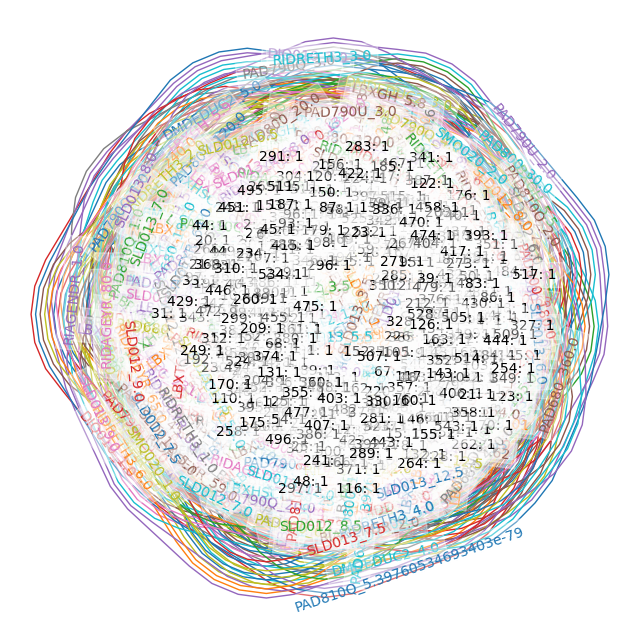

**3.2 Confusion matrices**

**Figure S3 –** Confusion matrices of the machine learning model built with the reduced feature sets

| 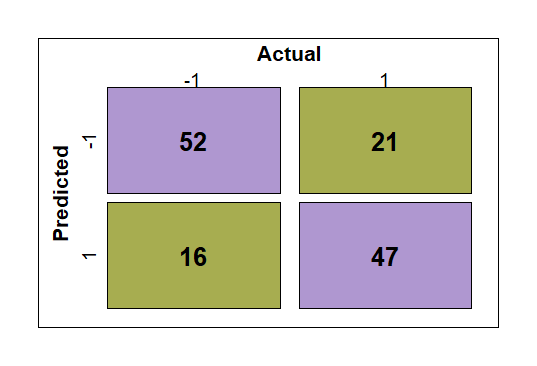   (a) **CVFE (*c* = 2, *e* = 10, *p* = 0.2)** | 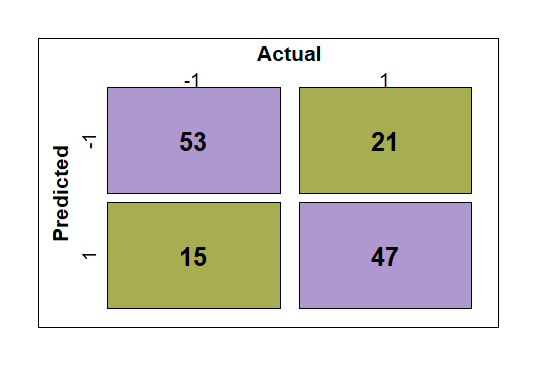   (b) **CVFE (*c* = 2, *e* = 10, *p* = 0.6)** | 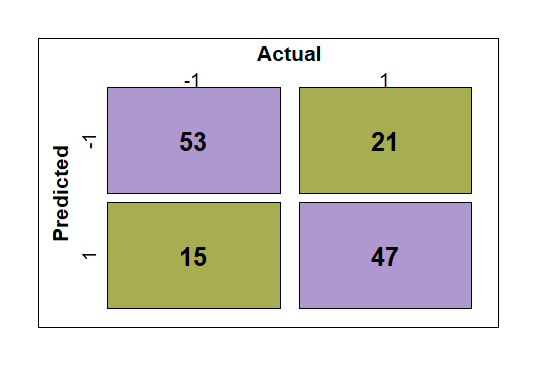   (c) **CVFE (*c* = 2, *e* = 5, *p* = 0.8)** | 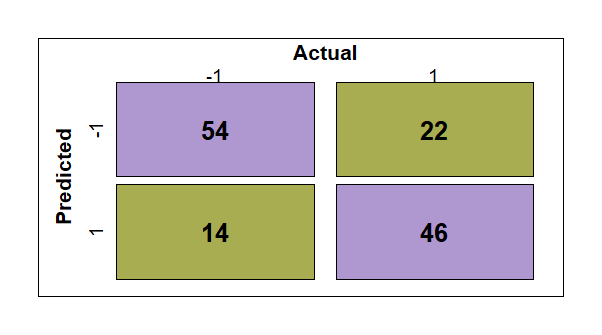   (d) **CVFE (*c* = 3, *e* = 5, *p* = 0.8)** |
| --- | --- | --- | --- |
| 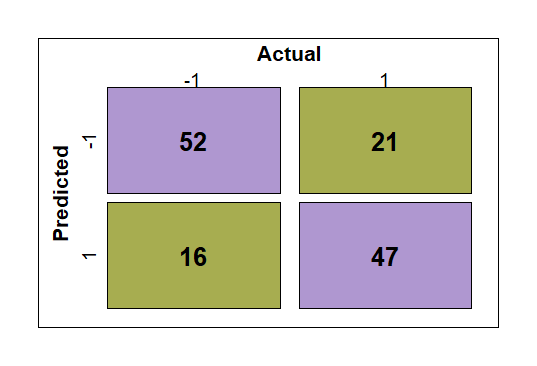   (e) **HFE (bin = 5, *β*** **= 25)** | 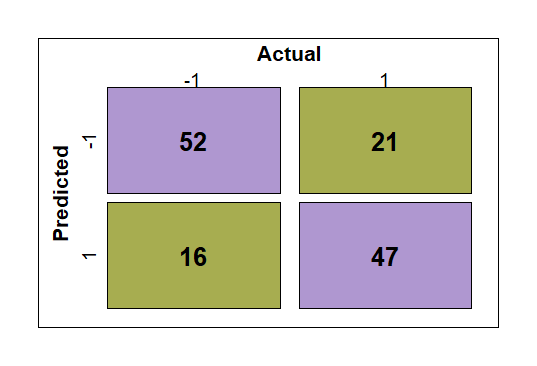   (f) **HFE (bin = 5, *β* = 50)** | 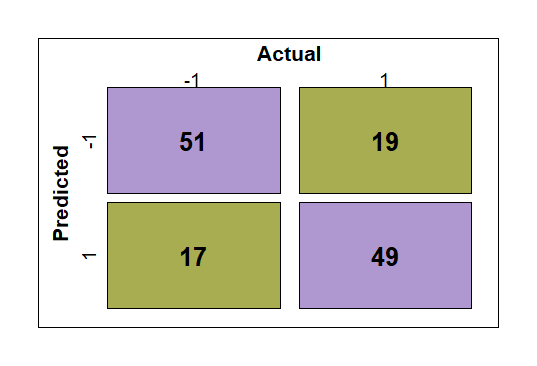   (g) **HFE (bin = 5, *β* = 75)** | 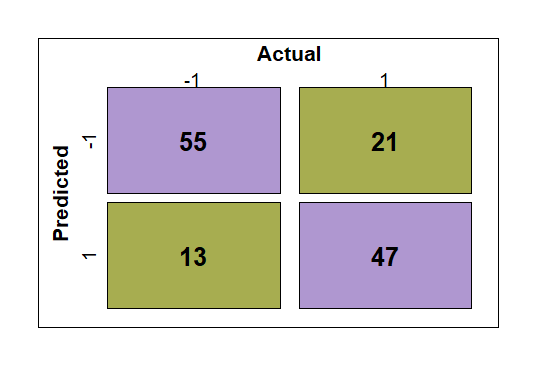   (h) **HFE (bin = 10, *β* = 25)** |
| 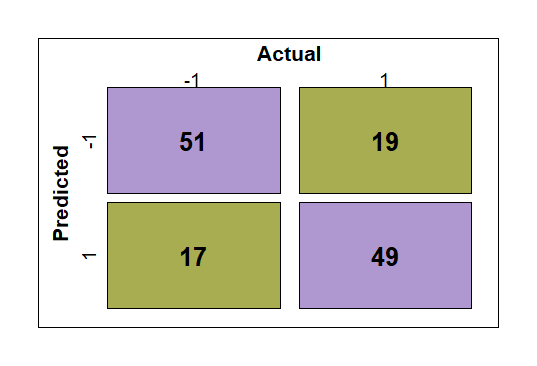   (i) **HFE (bin = 10, *β* = 50)** | 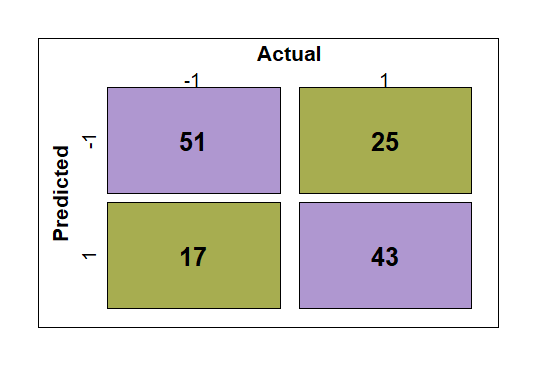   (j) **HFE (bin = 10, *β* = 75)** | 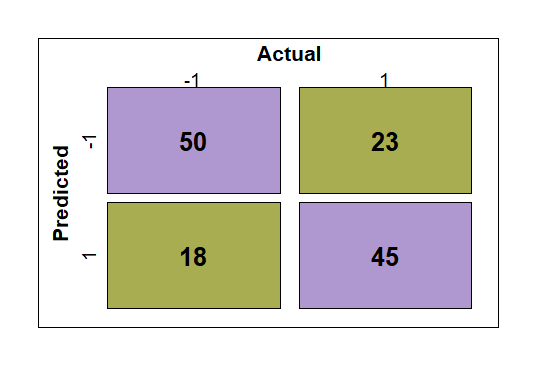   (k) **ADT** |  |
